# Comparative Effectiveness of mRNA-1273 and BNT162b2 COVID-19 Vaccines in Immunocompromised Individuals: A Systematic Review and Meta-Analysis Using the GRADE Framework

**DOI:** 10.1101/2023.04.05.23288195

**Authors:** Xuan Wang, Katrin Haeussler, Anne Spellman, Leslie E. Phillips, Allison Ramiller, Mary T. Bausch-Jurken, Pawana Sharma, Anna Krivelyova, Sonam Vats, Nicolas Van de Velde

**Affiliations:** ICON plc, Stockholm, Sweden; ICON plc, Munich, Germany; Data Health Ltd, London, United Kingdom; Data–Driven LLC, Seattle, WA, United States; Moderna, Inc., Cambridge, MA, United States; ICON plc, London, United Kingdom; ICON plc, Bengaluru, India

**Keywords:** Severe acute respiratory syndrome coronavirus 2, SARS-CoV-2, COVID-19, mRNA vaccine, mRNA-1273, BNT162b2, immunocompromised, effectiveness

## Abstract

**Introduction:** Despite representing only 3% of the US population, immunocompromised (IC) individuals account for nearly half of the COVID-19 breakthrough hospitalizations. IC individuals generate a lower immune response following vaccination in general, and the US CDC recommended a third dose of either mRNA-1273 or BNT162b2 COVID-19 vaccines as part of their primary series. Influenza vaccine trials have shown that increasing dosage could improve effectiveness in IC populations. The objective of this systematic literature review and pairwise meta-analysis was to evaluate the clinical effectiveness of mRNA-1273 (50 or 100 mcg/dose) versus BNT162b2 (30 mcg/dose) in IC populations using the GRADE framework.

**Methods:** The systematic literature search was conducted in the World Health Organization COVID-19 Research Database. Studies were included in the pairwise meta-analysis if they reported comparisons of mRNA-1273 and BNT162b2 in IC individuals ≥18 years of age; outcomes of interest were SARS-CoV-2 infection, hospitalization due to COVID-19, and mortality due to COVID-19. Risk ratios (RR) were pooled across studies using random-effects meta-analysis models. Outcomes were also analyzed in subgroups of patients with cancer, autoimmune disease, and solid organ transplant. Risk of bias was assessed for randomized and observational studies using the Risk of Bias 2 tool and the Newcastle-Ottawa Scale, respectively. Evidence was evaluated using the GRADE framework.

**Results:** Overall, 22 studies were included in the pairwise meta-analysis. Compared with BNT162b2, mRNA-1273 was associated with significantly reduced risk of SARS-CoV-2 infection (RR 0.87, 95% CI 0.79–0.96; *P*=0.0054; *I^2^*=61.9%), COVID-19–associated hospitalization (RR 0.83, 95% CI 0.76–0.90; *P*<0.0001; *I^2^*=0%), and COVID-19–associated mortality (RR 0.62, 95% CI 0.43–0.89; *P*=0.011; *I^2^*=0%) in IC populations. Results were consistent across subgroups. Because of sample size limitations, relative effectiveness of COVID-19 mRNA vaccines in IC populations cannot be studied in randomized trials and evidence certainty among comparisons was type 3 (low) and 4 (very low), reflecting potential biases in observational studies.

**Conclusion:** This GRADE meta-analysis based on a large number of consistent observational studies showed that the mRNA-1273 COVID-19 vaccine is associated with improved clinical effectiveness in IC populations compared with BNT162b2.

## Introduction

The global coronavirus disease 2019 (COVID-19) pandemic caused by severe acute respiratory syndrome coronavirus 2 (SARS-CoV-2) has resulted in 103 million reported infections and 1.1 million deaths to date in the United States (US) (1). In response to the pandemic, mRNA-1273 (Spikevax®, Moderna, Inc., Cambridge, USA) (2) and BNT162b2 (Comirnaty®, Pfizer/BioNTech, New York, USA/Mainz, Germany) (3), each employing novel mRNA technology, were developed and approved for the prevention of COVID-19 (4). Global phase 2/3 studies demonstrated that both mRNA vaccines given in a 2-dose series were highly efficacious at reducing symptomatic infections and hospitalizations in the immunocompetent population (5; 6).

Although immunocompromised (IC) individuals comprise only approximately 3% of the total US population (7), they account for nearly half of the breakthrough COVID-19 hospitalizations (8). While there is a range of severity across conditions at the population level, adults considered immunodeficient had 2.68-fold greater adjusted odds of being hospitalized with COVID-19 compared with immunocompetent individuals due both to the underlying IC condition and therapies used for treatment (9; 10). In 1 study, use of immunosuppression in patients with autoimmune disease resulted in 1.35-fold (95% confidence interval [CI] 1.29–1.40) greater odds of developing life-threatening COVID-19 (11).

Despite being at increased risk of COVID-19–related morbidity and mortality (10; 12-14), IC individuals and patients receiving immunosuppressive medications were excluded from participating in pivotal trials of mRNA-1273 and BNT162b2 (5; 6). Real-world COVID-19 data indicate that vaccine immune responses are generally impaired in IC populations (9; 15-17) and that vaccine effectiveness, as estimated as the odds of obtaining a positive SARS-CoV-2 test result using multivariate logistic regression models, is lower in IC versus immunocompetent individuals (18). In addition to severe COVID-19, IC populations are at higher risk of prolonged SARS-CoV-2 infection (19–26) and viral evolution (19-22; 24; 27; 28) due to poor humoral responses. These risks are exacerbated by even lower antibody responses to SARS-CoV-2 variants (29–35). IC individuals may also contribute disproportionately to SARS-CoV-2 transmission to household contacts (36). High vaccine effectiveness is therefore critically important for this population and the US Centers for Disease Control and Prevention (CDC) recommended a third dose of either mRNA-1273 or BNT162b2 COVID-19 vaccines as part of their primary series.

Influenza vaccine trials demonstrated that high dose vaccines led to improved immune responses in IC individuals compared with standard dose vaccines and suggested that a high dose vaccine offers greater effectiveness for IC populations (37–42). Although both mRNA-1273 and BNT162b2 employ the mRNA mode of action, the composition of each vaccine is different. For instance, the mRNA dosage and type of lipid nanoparticles used in the delivery system differs between vaccines. The mRNA-1273 primary series contains 100 mcg of mRNA and 50 mcg for the booster (2; 43), whereas BNT162b2 contains 30 mcg of mRNA for each primary and booster dose (3; 44). Observational studies have consistently shown differences between the two mRNA COVID-19 vaccines both in terms of immune response (15) and clinical effectiveness (45–47) in IC populations.

As SARS-CoV-2 transitions from a pandemic to an endemic state, countries are transferring vaccination programs from central government purchasing to their respective national healthcare systems, which is triggering in-depth health technology assessments to recommend the best use of available vaccines in specific populations. Several national immunization technical advisory groups (NITAGs), including the Advisory Committee on Immunization Practices (ACIP) in the US, use the GRADE (Grading of Recommendations, Assessment, Development and Evaluations) framework for identifying questions relevant to healthcare, selecting outcomes of interest and assessing their importance, evaluating the available evidence, and synthesizing evidence to develop recommendations consistent with considerations of values and preferences of patients and the society in which they live (48; 49).

This present analysis follows the GRADE framework to address the following healthcare question: Is the mRNA-1273 COVID-19 vaccine (50 or 100 mcg/dose) more clinically effective in IC populations compared with the BNT162b2 COVID-19 vaccine (30 mcg/dose)? Accordingly, we performed a systematic literature review and pairwise meta-analysis to compare COVID-19 vaccine effectiveness outcomes among IC individuals given either mRNA-1273 or BNT162b2.

## Methods

### Search strategy and study selection

We performed a systematic literature review in accordance with the Preferred Reporting Items for Systematic Reviews and Meta-Analyses 2020 framework (50). The main search was conducted in the World Health Organization COVID-19 Research Database on April 14, 2022 and updated on December 19, 2022. Databases searched were MEDLINE, International Clinical Trials Registry Platform, EMBASE, EuropePMC, medRxiv, Web of Science, ProQuest Central, Academic Search Complete, Scopus, and COVIDWHO. The search strategy is presented in **Table S1**.

**Table 1.** Characteristics of Studies Included in the Meta-Analysis.

| Author,<br>year | Study Characteristics |  |  |  |  |  | Outcomes Reported |  |  |
| --- | --- | --- | --- | --- | --- | --- | --- | --- | --- |
|  | Design | Data source | Population | Vaccine | Study period | Vaccinated, n | Infection | Hospitalization | Death |
| Aslam, 2021 (57) | • Retrospective single-center cohort | Transplant registry | • USA<br>• Solid organ transplant | • 2 doses (MM vs. PP)<br>• mRNA-1273 (100mcg)<br>• BNT162b2 (30mcg) | 01 Jan 2021 – 06 Feb 2021 | mRNA-1273: 632<br>BNT162b2: 375 | Y | N | Y |
| Britton, 2022 (58) | • Test-negative design | VISION Network, a collaboration between CDC and seven U.S. health care systems | • USA<br>• IC individuals | • 4 doses (MMMM vs. PPPP)<br>• mRNA-1273 (100mcg and booster 50mcg)<br>• BNT162b2 (30mcg) | Dec 2021 – Aug 2022 | mRNA-1273: 9,555<br>BNT162b2: 14,769 | Y | N | N |
| Butt, 2022 (59) | • Test-negative design<br>• 1:1 Propensity-score matched analysis of | Veterans Affairs | • USA<br>• Chronic hemodialysis | • 2 doses (MM vs. PP)<br>• mRNA-1273 | Jan 2021 – Aug 2021 | mRNA-1273: 630<br>BNT162b2: 719 | Y | N | N |
|  | cases and controls |  |  | (100mcg)<br>• BNT162b2 (30mcg) |  |  |  |  |  |
| Embi, 2021 (18) | • Test-negative design | VISION Network | • USA<br>• IC and immunocompetent vaccine recipients | • 2 doses (MM vs. PP)<br>• mRNA-1273 (100mcg)<br>• BNT162b2 (30mcg) |  | **IC and vaccinated**<br>mRNA-1273: 4,337<br>BNT162b2: 6,227<br>**Solid malignancy**<br>mRNA-1273: 2,053<br>BNT162b2: 2,848<br>**Rheumatologic or inflammatory disorder**<br>mRNA-1273: 1,053<br>BNT162b2: 1,591 | N | Y | N |
| Hause, 2022 (60) | • Health survey | VEARS | • USA<br>• “Presumed IC patients” as stated in the paper | • 4 doses (MMMM vs. PPPP)<br>• mRNA-1273 (100mcg) | v-safe Data: 12 Jan 2022–28 Mar 2022 | mRNA-1273: 2,194<br>BNT162b2: 1,821 | N | Y | N |
|  |  |  |  | and booster<br>50mcg)<br>• BNT162b2<br>(30mcg) |  |  |  |  |  |
| Holroyd,<br>2022 (61) | • Retrospective<br>single-center<br>study | CLIMB | • USA<br>• Patients with MS on disease modifying therapies vaccinated vs. healthy controls | • 2 doses (MM vs. PP)<br>• mRNA-1273 (100mcg)<br>• BNT162b2 (30mcg) | Jun 2021 – Dec 2021 | mRNA-1273: 110<br>BNT162b2: 133 | Y | N | N |
| Kelly, 2022 (62) | • Retrospective cohort study | US VHA | • USA<br>• IC patients including cancer | • mRNA-1273 (100mcg and booster 50mcg)<br>• BNT162b2 (30mcg) | Jul 2021 – May 2022 | mRNA-1273: 79,517<br>BNT162b2: 67,780 | Y | Y | N |
| Khan, 2021 (63) | • Retrospective cohort study | US VHA | • USA<br>• Patients with inflammatory bowel disease exposed to various conventional and advanced immunosuppressive therapies | • 1 or 2 doses (MM vs. PP)<br>• mRNA-1273 (100mcg)<br>• BNT162b2 (30mcg) | 18 Dec 2020 (index date) – 20 Apr 2021 | **Fully vaccinated**<br>mRNA-1273: n=3,380<br>BNT162b2: n=2,873 | Y | N | N |
| Liew, 2022 (73) | Retrospective registry study | REDCap | Multi-country with 65% of the patient population from North America<br>Rheumatic disease | 2 doses (MM vs. PP)<br>mRNA-1273 (100mcg)<br>BNT162b2 (30mcg) | Jan 2021 – Sep 2021 | mRNA-1273: 45<br>BNT162b2: 21 | N | Y | N |
| Malinis, 2021 (68) | Retrospective observational study | Yale New Haven chart review | USA<br>Solid organ transplant recipients | 2 doses (MM vs PP)<br>mRNA-1273 (100 mcg)<br>BNT162b2 (30 mcg) | As of May 18, 2011 (start date not reported) | mRNA-1273: 157<br>BNT162b2: 275 | Y | N | N |
| Mazuecos, 2022 (46) | Retrospective national cohort study | National registry of patients with kidney transplantation | Spain<br>Kidney transplant recipients | 2 doses (MM vs. PP)<br>mRNA-1273 (100mcg)<br>BNT162b2 (30mcg) | Apr 2021 – Oct 2021 | mRNA-1273: 213<br>BNT162b2: 121 | N | Y | Y |
| Mues, 2022 (45) | Observational comparative effectiveness study<br>1:1 propensity- |  | USA<br>IC individuals | 2 doses (MM vs. PP)<br>mRNA-1273 | 11 Dec 2020 – 10 Jan 2022 | **IC**<br>mRNA-1273: 57,000<br>BNT162b2: 66,757 | Y | Y | N |
|  | score matched on age, sex, payer type, state of residence, previous healthcare utilization, comorbidities, frailty score and immunocompromised group |  |  | (100mcg)<br>• BNT162b2 (30mcg) |  | **Solid organ transplant**<br>mRNA-1273: 4,029<br>BNT162b2: 5,043<br>**Active cancer**<br>mRNA-1273: 7,186<br>BNT162b2: 8,277 |  |  |  |
| Patel, 2022 (47) | • Retrospective cohort study | MGB healthcare system | • USA<br>• Rheumatic disease | • ≥2 doses (MMM vs. PPP)<br>• mRNA-1273 (100mcg and booster 50mcg)<br>• BNT162b2 (30mcg) | Nov 2021 – Dec 2022 | mRNA-1273: 4,588<br>BNT162b2: 6,080 | Y | N | N |
| Pham, 2022 (64) | • Retrospective cohort study |  | • USA<br>• Primary immunodeficiency patients with functional B-cell defects | • 2 doses (MM vs. PP)<br>• mRNA-1273 (100mcg)<br>• BNT162b2 |  | mRNA-1273: 10<br>BNT162b2: 23 | Y | N | N |
|  |  |  |  | (30mcg) |  |  |  |  |  |
| Piñana, 2022 (69) | Prospective multicenter registry-based cohort study | GRUCINI with SEHH | Spain<br>Patients with hematological disorders | 2 doses (MM vs. PP)<br>mRNA-1273 (100mcg)<br>BNT162b2 (30mcg) | Dec 2020 – Dec 2021 | mRNA-1273: 982<br>BNT162b2: 362 | Y | N | N |
| Pino, 2022 (70) | Retrospective cohort study | Patients followed at the Medical Oncology Unit in Florence at Santa Maria Annunziata, Serristori and Borgo San Lorenzo Hospitals | Italy<br>Extremely vulnerable individuals, patients with cancer on systemic antitumor treatment | 3 doses (MMM vs. PPP)<br>mRNA-1273 (100mcg and booster 50mcg)<br>BNT162b2 (30mcg) | 26 Mar 2021 – 04 Apr 2021 | mRNA-1273: 527<br>BNT162b2: 96 | Y | N | N |
| Quiroga, 2022 (32) | Prospective real-world study | SENCOVAC (multicentric study by the Spanish Society of Nephrology) | Spain<br>Patients on hemodialysis | Booster dose (primary vaccination BNT162b2 or mRNA-1273 or |  | mRNA-1273: 481<br>BNT162b2: 230 | Y | N | N |
|  |  |  |  | ChAdOx1-S)<br>• mRNA-1273 (booster 50mcg)<br>• BNT162b2 (30mcg) |  |  |  |  |  |
| Rooney, 2022 (65) | • Retrospective study | The University of Kansas Cancer Center Curated Cancer Clinical Outcomes Database was queried | • USA<br>• Patients with cancer receiving antineoplastic therapy | • 2 doses (MM vs. PP)<br>• mRNA-1273 (100mcg)<br>• BNT162b2 (30mcg) | Feb 2021 – Oct 2021 | mRNA-1273: 2,993<br>BNT162b2: 6,423 | Y | N | N |
| Sibbel, 2021 (66) | • Retrospective observational study |  | • USA<br>• Hemodialysis patients | • 1 or 2 doses (MM vs. PP)<br>• mRNA-1273 (100mcg)<br>• BNT162b2 (30mcg) | Study duration: 4 months | mRNA-1273: 23,037<br>BNT162b2: 12,169 | Y | Y | Y |
| Speich, | • Parallel, 2-arm | Swiss HIV | • Switzerland | • 2 doses |  | mRNA-1273: | Y* | N | N |
| 2022 (72) | (allocation 1:1), open-label, noninferiority RCT nested into the Swiss HIV Cohort Study and the Swiss Transplant Cohort Study, all outcomes were assessed 12 weeks ($\pm 7$ days) after the first vaccination.<br>• Cohorts were stratified by study center, age group, sex, and presence of comorbidities | Cohort, Swiss Transplant Cohort | • IC individuals (HIV and transplant) | (MM vs. PP)<br>• mRNA-1273 (100mcg)<br>• BNT162b2 (30mcg) | | 205<br>BNT162b2: 210 | | | |
| Yeo, 2022 (71) | • Prospective, observational study |  | • Singapore<br>• Patients with MS, AQP4-NMOSD, and MOGAD | • 2-3 doses (MMM vs. PPP)<br>• mRNA-1273 (100mcg and booster | Study cut-off date: 31 Dec 2022 | mRNA-1273: 38<br>BNT162b2: 327 | Y | Y | N |
|  |  |  |  | 50mcg)<br>• BNT162b2<br>(30mcg) |  |  |  |  |  |
| Yetmar,<br>2022 (67) | • Retrospective<br>cohort | From 1 US<br>center | • USA<br>• Solid organ<br>transplant<br>recipients | • 2 doses<br>(MM vs.<br>PP)<br>• mRNA-1273<br>(100mcg)<br>• BNT162b2<br>(30mcg) | Aug 2021 –<br>Sep 2021 | mRNA-1273:<br>12<br>BNT162b2:<br>22 | Y | N | Y |

Vaccine dosing was abbreviated as 'MM,' 'MMM,' or 'MMMM' for 2, 3, or 4 doses of mRNA-1273, respectively, and as 'PP,' 'PPP,' or 'PPPP' for 2, 3, or 4 doses of BNT162b2, respectively. \* Laboratory-confirmed, symptomatic infection. AQP4-NMSOD, aquaporin-4-antibody neuromyelitis optica spectrum disorder; CDC, Centers for Disease Control and Prevention; CLIMB, Comprehensive Longitudinal Investigation of Multiple Sclerosis at Brigham and Women's Hospital; GRUCINI, Infectious Complications Subcommittee of the Spanish Hematopoietic Stem Cell Transplantation and Cell Therapy Group; HIV, human immunodeficiency virus; IC, immunocompromised; MGB, Mass General Brigham; MOGAD, myelin oligodendrocyte glycoprotein-antibody associated disease; MS, multiple sclerosis; SEHH, Spanish Society of Hematology and Hemotherapy; VAERS, Vaccine Adverse Event Reporting System; VHA, Veterans Health Administration.

Clinical trials, observational studies, or any real-world evidence published as manuscripts, letters, commentaries, abstracts, or posters were included if they reported efficacy or clinical effectiveness outcomes in IC individuals ≥18 years of age vaccinated with mRNA-1273 or BNT162b2 within the same study. IC individuals were defined as people with immunocompromising conditions falling into the clinically extremely vulnerable (CEV) groups 1 or 2, which include solid organ transplant, solid and hematological cancers, hemodialysis, poorly controlled human immunodeficiency virus (HIV) infection, and autoimmune conditions requiring immunosuppressive therapy (51). Outcomes of interest were vaccine efficacy or effectiveness against SARS-CoV-2 infection, COVID-19–associated hospitalization, and COVID-19– associated death. Recently published systematic literature reviews on the same topic were cross-checked to ensure relevant articles were included. Studies reporting outcomes in pregnant women, current or former smokers, or physically inactive people; with heterologous vaccination schedule (i.e., mix of mRNA-1273 and BNT162b2); with only safety data; or study protocols or economic models were excluded (**Table S2**). Two independent reviewers selected studies following a two-level approach; a third reviewer arbitrated conflicts. Titles and abstracts were screened against inclusion criteria in level 1, followed by full-text appraisal of articles not excluded at level 1 against selection criteria in level 2.

**Table 2.**
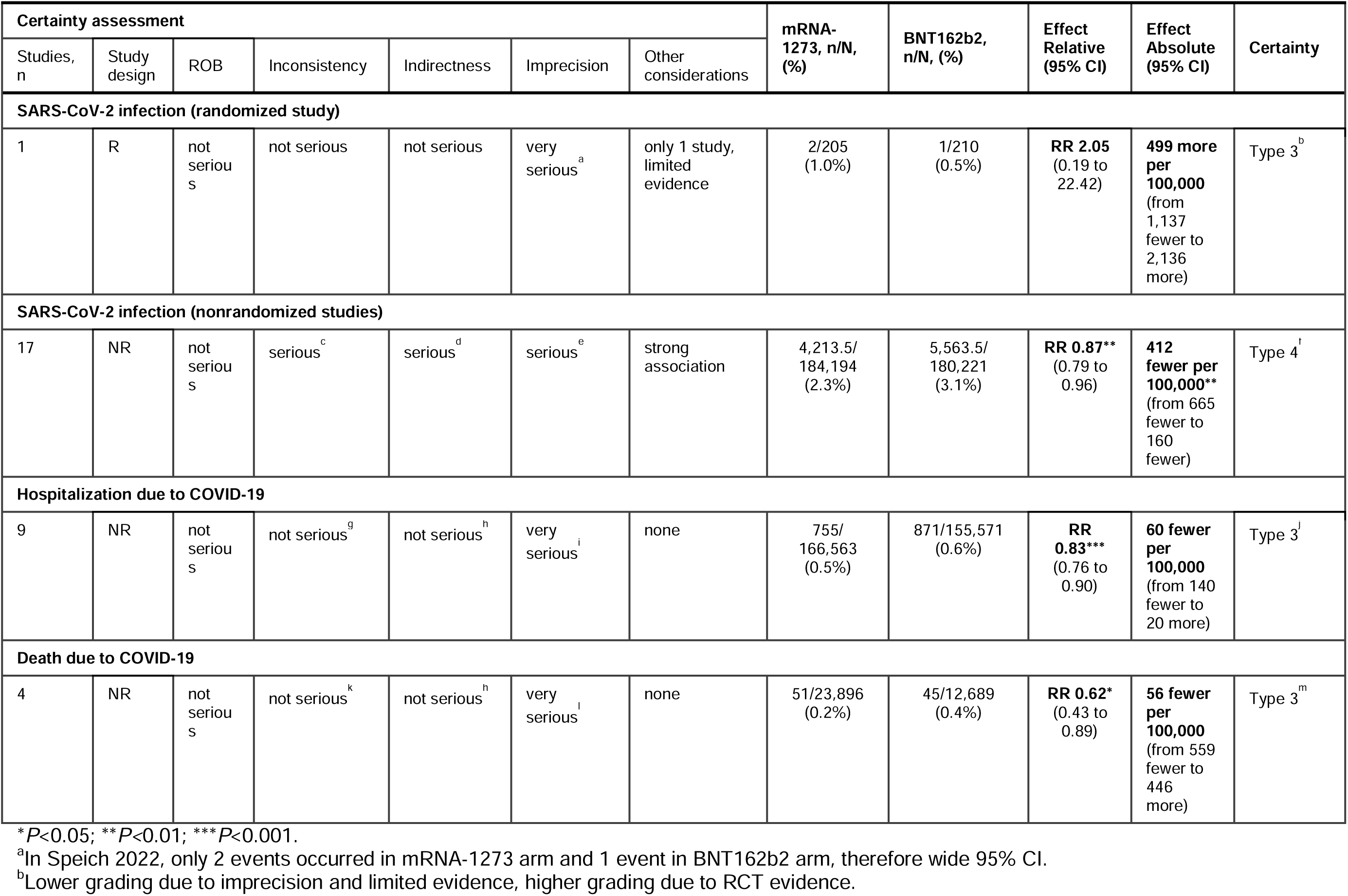
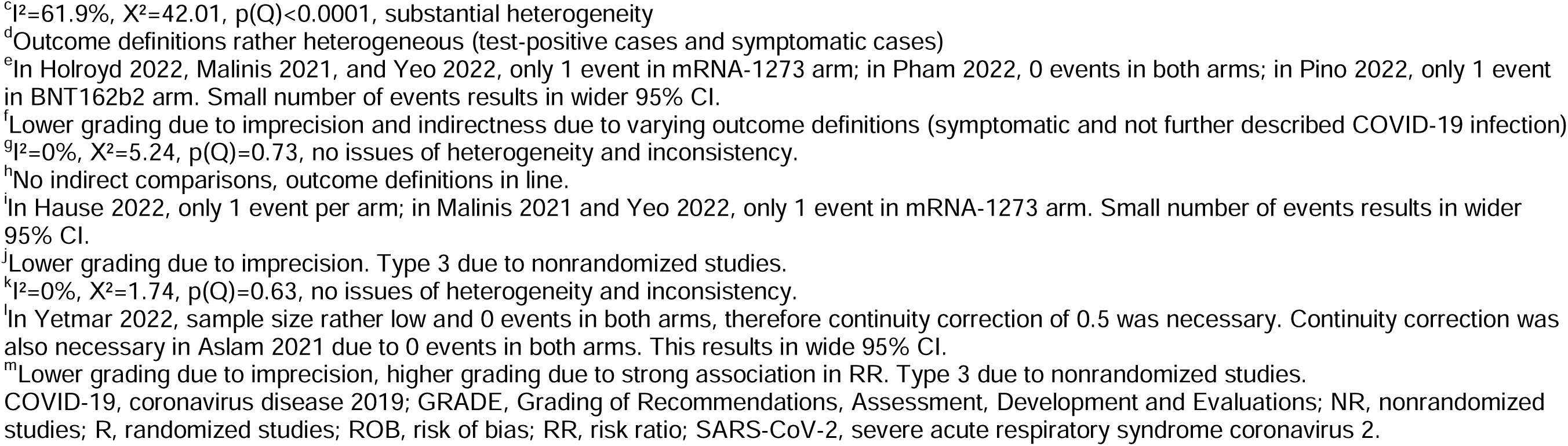
GRADE Summary of Findings Overall.

| Certainty assessment |  |  |  |  |  |  | mRNA-1273, n/N, (%) | BNT162b2, n/N, (%) | Effect Relative (95% CI) | Effect Absolute (95% CI) | Certainty |
| --- | --- | --- | --- | --- | --- | --- | --- | --- | --- | --- | --- |
| Studies, n | Study design | ROB | Inconsistency | Indirectness | Imprecision | Other considerations |  |  |  |  |  |
| SARS-CoV-2 infection (randomized study) |  |  |  |  |  |  |  |  |  |  |  |
| 1 | R | not serious | not serious | not serious | very serious<sup>a</sup> | only 1 study, limited evidence | 2/205 (1.0%) | 1/210 (0.5%) | RR 2.05 (0.19 to 22.42) | 499 more per 100,000 (from 1,137 fewer to 2,136 more) | Type 3<sup>b</sup> |
| SARS-CoV-2 infection (nonrandomized studies) |  |  |  |  |  |  |  |  |  |  |  |
| 17 | NR | not serious | serious<sup>c</sup> | serious<sup>d</sup> | serious<sup>e</sup> | strong association | 4,213.5/184,194 (2.3%) | 5,563.5/180,221 (3.1%) | RR 0.87** (0.79 to 0.96) | 412 fewer per 100,000** (from 665 fewer to 160 fewer) | Type 4<sup>f</sup> |
| Hospitalization due to COVID-19 |  |  |  |  |  |  |  |  |  |  |  |
| 9 | NR | not serious | not serious<sup>g</sup> | not serious<sup>h</sup> | very serious<sup>i</sup> | none | 755/166,563 (0.5%) | 871/155,571 (0.6%) | RR 0.83*** (0.76 to 0.90) | 60 fewer per 100,000 (from 140 fewer to 20 more) | Type 3<sup>j</sup> |
| Death due to COVID-19 |  |  |  |  |  |  |  |  |  |  |  |
| 4 | NR | not serious | not serious<sup>k</sup> | not serious<sup>h</sup> | very serious<sup>l</sup> | none | 51/23,896 (0.2%) | 45/12,689 (0.4%) | RR 0.62* (0.43 to 0.89) | 56 fewer per 100,000 (from 559 fewer to 446 more) | Type 3<sup>m</sup> |

\* $P < 0.05$ ; \*\* $P < 0.01$ ; \*\*\* $P < 0.001$ .
<sup>a</sup>In Speich 2022, only 2 events occurred in mRNA-1273 arm and 1 event in BNT162b2 arm, therefore wide 95% CI.
<sup>b</sup>Lower grading due to imprecision and limited evidence, higher grading due to RCT evidence.
<sup>c</sup> $I^2=61.9\%$ , $X^2=42.01$ , $p(Q)<0.0001$ , substantial heterogeneity
<sup>d</sup>Outcome definitions rather heterogeneous (test-positive cases and symptomatic cases)
<sup>e</sup>In Holroyd 2022, Malinis 2021, and Yeo 2022, only 1 event in mRNA-1273 arm; in Pham 2022, 0 events in both arms; in Pino 2022, only 1 event in BNT162b2 arm. Small number of events results in wider 95% CI.
<sup>f</sup>Lower grading due to imprecision and indirectness due to varying outcome definitions (symptomatic and not further described COVID-19 infection)
<sup>g</sup> $I^2=0\%$ , $X^2=5.24$ , $p(Q)=0.73$ , no issues of heterogeneity and inconsistency.
<sup>h</sup>No indirect comparisons, outcome definitions in line.
<sup>i</sup>In Hause 2022, only 1 event per arm; in Malinis 2021 and Yeo 2022, only 1 event in mRNA-1273 arm. Small number of events results in wider 95% CI.
<sup>j</sup>Lower grading due to imprecision. Type 3 due to nonrandomized studies.
<sup>k</sup> $I^2=0\%$ , $X^2=1.74$ , $p(Q)=0.63$ , no issues of heterogeneity and inconsistency.
<sup>l</sup>In Yetmar 2022, sample size rather low and 0 events in both arms, therefore continuity correction of 0.5 was necessary. Continuity correction was also necessary in Aslam 2021 due to 0 events in both arms. This results in wide 95% CI.
<sup>m</sup>Lower grading due to imprecision, higher grading due to strong association in RR. Type 3 due to nonrandomized studies.

### Data extraction and quality assessment

Publication details, study and participant characteristics, vaccine type and vaccination status, at-risk condition, and clinical outcomes were extracted. Risk of bias was assessed in accordance with Cochrane review guidelines (52) using the ROB 2 tool (53) for randomized studies and the Newcastle-Ottawa Scale (54) for observational studies. Evidence was evaluated based on GRADE criteria (48; 49).

### Statistical analysis

Random-effects meta-analysis models were used to pool risk ratios (RR) and calculate absolute effects as risk difference (RD) per 100,000 individuals across studies. Inverse variance weights were calculated for individual studies with the DerSimonian-Laird method (55). Chi-square testing to evaluate heterogeneity across studies was performed (56). The *I^2^*statistic was estimated (0–100%, 0% meaning no evidence of heterogeneity). Subgroup analysis was performed for patients with cancer, autoimmune disease, and solid organ transplant.

## Results

### Overview of included studies

Of 5,745 unique items retrieved, we identified 35 studies reporting COVID-19 clinical efficacy or effectiveness outcomes in IC individuals ≥18 years of age who received mRNA-1273 or BNT162b2 in the same study (**Figure 1**). Thirteen articles were excluded because the population did not meet the inclusion criteria (i.e., participants had immunocompromising conditions not included in CEV groups 1 or 2), 1-dose vaccine regimen data were reported, or the outcome of interest data were not reported in sufficient detail for analysis. Characteristics of all nonrandomized (n=21) and randomized (n=1) studies included in the pairwise meta-analysis are shown in **Table 1**. Overall, 190,643 and 187,813 patients received mRNA-1273 and BNT162b2, respectively. Studies included mostly US populations (n=15) (18; 45; 47; 57-68), with the remaining trials reporting data on patients from Spain (n=3) (32; 46; 69), Italy (n=1) (70), Singapore (n=1) (71), Switzerland (n=1) (72), and multiple countries (n=1) (73). Specific at-risk and IC conditions included solid organ transplant (n=6) (45; 46; 57; 67; 68; 72), cancer (n=5) (18; 45; 62; 65; 70), hemodialysis (n=3) (32; 59; 66), rheumatologic disease (n=3) (18; 47; 73), multiple sclerosis or other neurological autoimmune disease (n=2) (61; 71), inflammatory bowel disease (n=1) (63), primary immunodeficiency with functional B cell defects (n=1) (64), hematological disorders (n=1) (69), and HIV infection (n=1) (72). Two studies did not specify the IC condition (58; 60). Individuals received ≥2 doses of mRNA-1273 or BNT162b2. Data on 2-dose regimens were considered if reported (n=15) (18; 45; 46; 57; 59; 61; 63-69; 72; 73), otherwise data from 3-or 4-dose regimens (n=7) (32; 47; 58; 60; 62; 70; 71) were used. Of studies reporting data from 2-dose regimens, outcomes were assessed ≥14 days after the second dose (n=13), ≥7 days after the second dose (n=1) (63), and other timepoints (n=1) (66). Timing of outcome assessment relative to the second dose was not specified in 3 studies. Variants of concern were delta (n=6) (18; 45; 46; 65; 67; 69), delta and omicron (n=2) (62; 71), delta and beta (n=1) (59), pre-omicron variants (n=1) (47), and omicron only (n=1) (58). Eleven studies did not directly specify the variant assessed (32; 57; 60; 61; 63; 64; 66; 68; 70; 72; 73).

**Figure 1.**
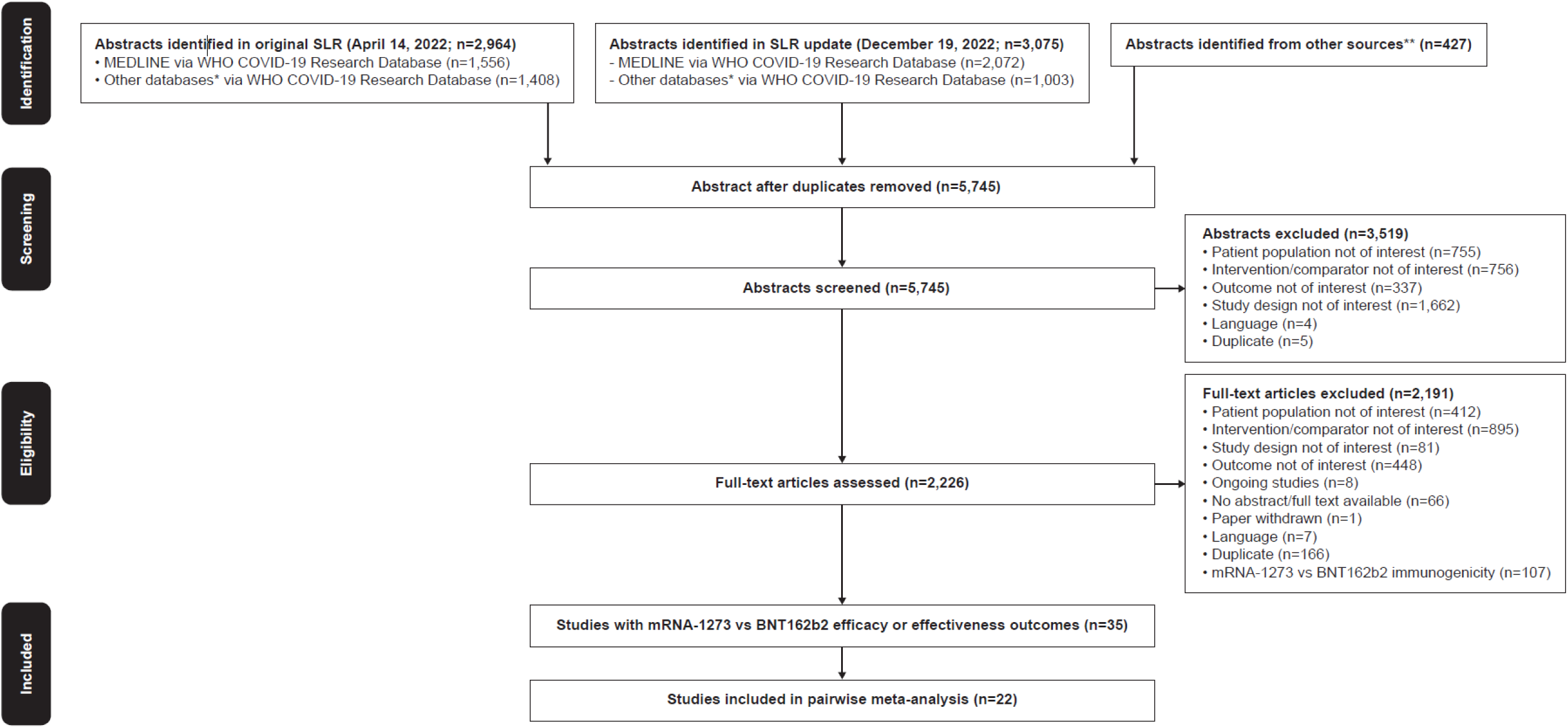
PRISMA Flow Diagram. Searches were first performed on April 14, 2022 followed by an update on December 19, 2022. * Databases searched include ICTRP, EMBASE, EuropePMC, medRxiv, Web of Science, ProQuest Central, Academic Search Complete, Scopus, and COVIDWHO. ** Includes internal documents from Moderna and recently published SLRs. COVID-19, coronavirus disease 2019; PRISMA, Preferred Reporting Items for Systematic Reviews and Meta-Analyses; SLR, systematic literature review; WHO, World Health Organization.

Risk of bias assessment found no serious risk of bias for 17 studies (randomized, n=1; nonrandomized, n=16) and serious risk of bias in 5 nonrandomized studies primarily due to the lack of description of comparability between cohorts or adjustment for confounding factors (**Table S3**; **Table S4**).

**Table 3.** GRADE Summary of Findings by Population Subgroup.

| Certainty assessment |  |  |  |  |  |  | mRNA<br>-1273,<br>n/N,<br>(%) | BNT162b2,<br>n/N, (%) | Effect<br>Relative<br>(95% CI) | Effect<br>Absolute<br>(95% CI) | Certainty |
| --- | --- | --- | --- | --- | --- | --- | --- | --- | --- | --- | --- |
| Studies,<br>n | Study<br>design | ROB | Inconsistency | Indirectness | Imprecision | Other<br>considerations |  |  |  |  |  |
| SARS-CoV-2 infection |  |  |  |  |  |  |  |  |  |  |  |
| Solid organ transplant |  |  |  |  |  |  |  |  |  |  |  |
| 4 | R | not<br>seriou<br>s | not serious<sup>a</sup> | serious<sup>e</sup> | very<br>serious<sup>b</sup> | none | 106/<br>4,830<br>(2.2%) | 139/5,715<br>(2.4%) | RR 1.05<br>(0.87 to<br>1.26) | 93 fewer per<br>100,000<br>(from 573<br>fewer to 386<br>more) | Type 4<sup>c</sup> |
| Cancer |  |  |  |  |  |  |  |  |  |  |  |
| 4 | NR | not<br>seriou<br>s | not serious<sup>d</sup> | serious<sup>e</sup> | very serious<sup>i</sup> | strong<br>association | 116/<br>11,688<br>(1.0%) | 190/15,158<br>(1.3%) | RR 0.73**<br>(0.57 to<br>0.92) | 346 fewer<br>per 100,000**<br>(from 598<br>fewer to 94<br>fewer) | Type 4<sup>c</sup> |
| Autoimmune disease |  |  |  |  |  |  |  |  |  |  |  |
| 4 | NR | not<br>seriou<br>s | not serious<sup>g</sup> | serious<sup>e</sup> | very<br>serious<sup>h</sup> | strong<br>association | 83/<br>8,116<br>(1.0%) | 176/9,413<br>(1.9%) | RR 0.67**<br>(0.52 to<br>0.88) | 455 fewer<br>per 100,000<br>(from 1,209<br>fewer to 298<br>fewer) | Type 4<sup>c</sup> |
| Hospitalization due to COVID-19 |  |  |  |  |  |  |  |  |  |  |  |
| Solid organ transplant |  |  |  |  |  |  |  |  |  |  |  |
| 3 | NR | not<br>seriou<br>s | not serious<sup>i</sup> | not serious<sup>j</sup> | not serious<sup>k</sup> | none | 163/<br>4,448<br>(3.7%) | 123/5,488<br>(2.2%) | RR 0.91<br>(0.78 to<br>1.06) | 147 fewer<br>per 100,000<br>(from 816<br>fewer to 522<br>more) | Type 3<sup>i</sup> |
| Cancer |  |  |  |  |  |  |  |  |  |  |  |
| 2 | NR | not<br>seriou<br>s | not serious<sup>m</sup> | not serious<sup>j</sup> | very<br>serious<sup>n</sup> | none | 38/<br>9,239<br>(0.4%) | 94/11,125<br>(0.8%) | RR 0.54**<br>(0.37 to<br>0.79) | 585 fewer<br>per 100,000<br>(from 1,655<br>fewer to 485<br>more) | Type 3<sup>i</sup> |

| Autoimmune disease |  |  |  |  |  |  |  |  |  |  |  |
| --- | --- | --- | --- | --- | --- | --- | --- | --- | --- | --- | --- |
| 3 | NR | not serious | not serious<sup>o</sup> | not serious<sup>j</sup> | very serious<sup>p</sup> | none | 54/1,112 (4.9%) | 93/1,963 (4.7%) | RR 0.97 (0.70 to 1.35) | 103 fewer per 100,000 (from 1,661 fewer to 1,456 more) | Type 3<sup>i</sup> |
| Death due to COVID-19 |  |  |  |  |  |  |  |  |  |  |  |
| Solid organ transplant |  |  |  |  |  |  |  |  |  |  |  |
| 3 | NR | not serious | not serious<sup>q</sup> | not serious<sup>j</sup> | very serious<sup>r</sup> | strong association | 36/859 (4.2%) | 37/520 (7.1%) | RR 0.56** (0.37 to 0.84) | 3,528 fewer per 100,000 (from 12,002 fewer to 4,945 more) | Type 3<sup>s</sup> |

\* $P < 0.05$ ; \*\* $P < 0.01$ ; \*\*\* $P < 0.001$ .
<sup>a</sup> $I^2 = 0\%$ , $X^2 = 0.78$ , $p(Q) = 0.61$ , no issues of heterogeneity and inconsistency.
<sup>b</sup>In Aslam 2021, only 2 events in mRNA-1273 and BNT162b2 arms; in Malinis 2021, only 1 event in mRNA-1273 and 2 events in the BNT162b2 arms. Small number of events results in a wide 95% CI.
<sup>c</sup>Lower grading due to imprecision, limited evidence and indirectness due to varying outcome definitions (symptomatic and not further described COVID-19 infection)
<sup>d</sup> $I^2 = 0\%$ , $X^2 = 0.13$ , $p(Q) = 0.99$ , no issues of heterogeneity and inconsistency.
<sup>e</sup>Outcome definitions rather heterogeneous (test-positive cases and symptomatic cases)
<sup>f</sup>In Pino 2022, only 1 event in BNT162b2 arm; this results in wider 95% CI.
<sup>g</sup> $I^2 = 0\%$ , $X^2 = 0.78$ , $p(Q) = 0.85$ , no issues of heterogeneity and inconsistency.
<sup>h</sup>In Holroyd 2022 and Yeo 2022, only 1 event in mRNA-1273 arm; this results in wider 95% CI.
<sup>i</sup> $I^2 = 0\%$ , $X^2 = 0.71$ , $p(Q) = 0.70$ , no issues of heterogeneity and inconsistency.
<sup>j</sup>No indirect comparisons, outcome definitions in line.
<sup>k</sup>No issues with low numbers of events apart from 1 event in the mRNA-1273 arm of Malinis 2021.
<sup>l</sup>Lower grading due to limited evidence. Type 3 due to nonrandomized studies.
<sup>m</sup> $I^2 = 0\%$ , $X^2 = 0.02$ , $p(Q) = 0.89$ , no issues of heterogeneity and inconsistency.

**Table 4.** Summary of Evidence for Outcomes of Interest.

| **Outcome** | **Outcome Importance<sup>a</sup>** | **Included in evidence profile** | **Certainty** |
| --- | --- | --- | --- |
| SARS-CoV-2 infection (randomized study) | Limited evidence | Yes | Type 3 (low) |
| SARS-CoV-2 infection (nonrandomized studies) | Critical | Yes | Type 4 (very low) |
| Hospitalization due to COVID-19 | Critical | Yes | Type 3 (low) |
| Death due to COVID-19 | Critical | Yes | Type 3 (low) |

<sup>a</sup> Relative importance of outcomes assessed and ranked per the GRADE framework. COVID-19, coronavirus disease 2019; GRADE, Grading of Recommendations, Assessment, Development, and Evaluations; SARS-CoV-2, severe acute respiratory syndrome coronavirus 2.

### SARS-CoV-2 infection (randomized study)

Only 1 and 2 laboratory-confirmed, symptomatic SARS-CoV-2 infections occurred in the BNT162b2 and mRNA-1273 arms, respectively, of a single randomized controlled trial (RCT; RR 2.05, 95% CI 0.19–22.42; RD 499, 95% CI −1,137 to 2,136) (72). The small number of events led to uncertainty around the estimates of effect and no association between mRNA vaccine type and risk of SARS-CoV-2 infection was found in this RCT. Evidence certainty was downgraded from type 1 (high) to type 3 (low) for imprecision and limited evidence (**Table 2; Table S4**).

### SARS-CoV-2 infection (nonrandomized studies)

Of the 17 nonrandomized studies reporting SARS-CoV-2 infection, mRNA-1273 was associated with a statistically significant reduction in the risk of SARS-CoV-2 infection compared with BNT162b2 (RR 0.87, 95% CI 0.79–0.96; *P*=0.0054; **Figure 2**). The RD (95% CI) of mRNA-1273 versus BNT162b2 was estimated to be 412 fewer SARS-CoV-2 infections (from 665 fewer to 160 fewer). Heterogeneity between studies may be considered substantial (*I^2^*=61.9%). The certainty of evidence was graded as type 4 (very low) for imprecision and indirectness due to varying outcome definitions (**Table 2; Table S3**).

**Figure 2.**
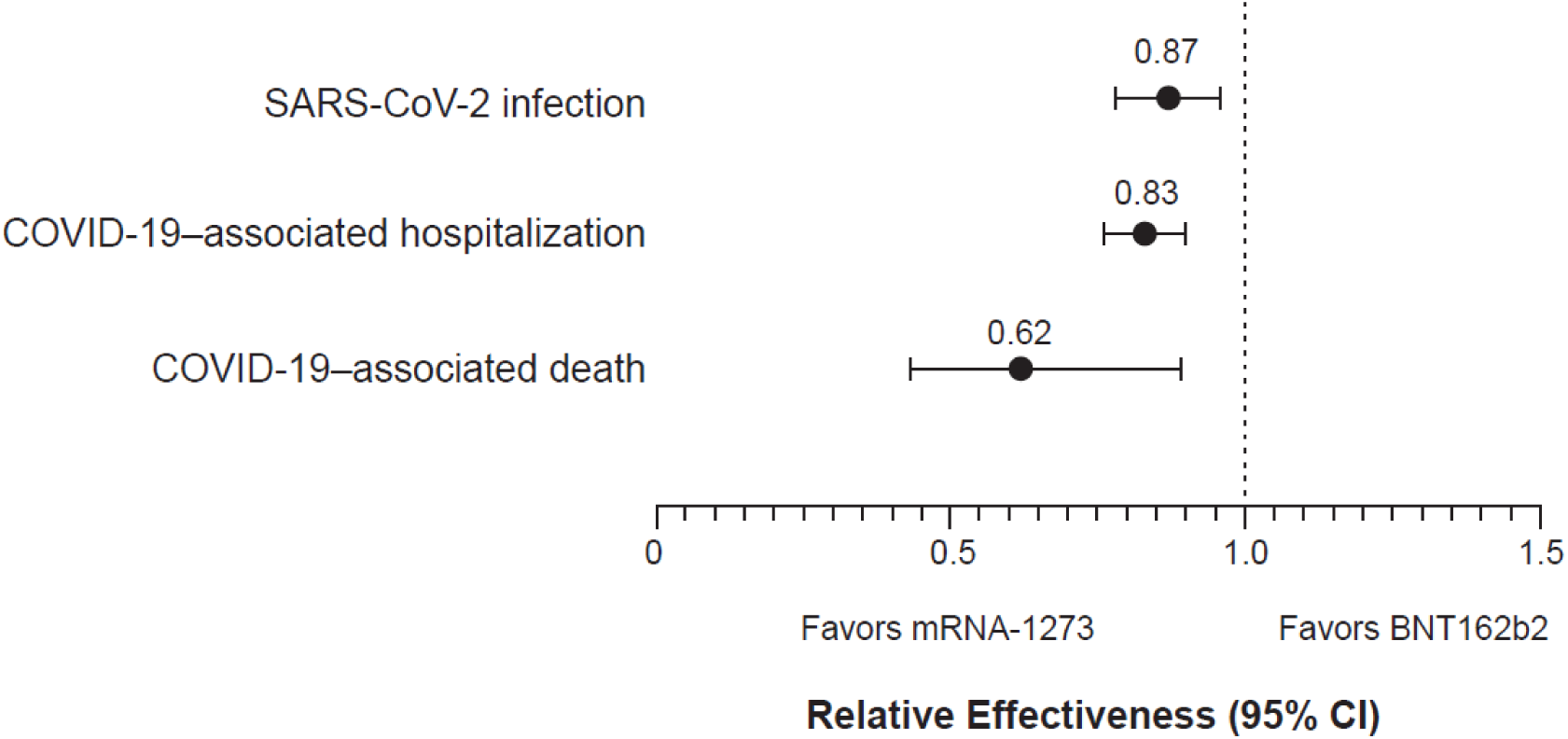
Summary of the Clinical Effectiveness Meta-Analysis. Relative risks of clinical effectiveness outcomes for mRNA-1273 versus BNT162b2 COVID-19 vaccines in IC populations are shown. CI, confidence interval; COVID-19, coronavirus disease 2019; IC, immunocompromised; SARS-CoV-2, severe acute respiratory syndrome coronavirus 2.

Analysis of 4 studies each reporting SARS-CoV-2 infection in patients with cancer (45; 65; 69; 70) found that mRNA-1273 was associated with significantly reduced risk of infection compared with BNT162b2 (RR 0.73, 95% CI 0.57–0.92, *P*=0.0088; RD −346, 95% CI −598 to −94, *P*=0.0071). Similar findings were observed in 4 studies assessing patients with autoimmune diseases (RR 0.67, 95% CI 0.52–0.88; *P*=0.0032; RD −455, 95% CI −1,209 to 298) (47; 61; 63; 71). No association between mRNA vaccine type was found for the 4 studies reporting SARS-CoV-2 infection in solid organ transplant recipients (RR 1.05, 95% CI 0.87– 1.26; RD −93, 95% CI −573 to 386) (45; 57; 67; 68). No evidence of heterogeneity was observed between any of the studies (*I^2^*=0% for all subgroups). As in the overall analysis of SARS-CoV-2 infection, the certainty of evidence was graded as type 4 (very low; **Table 3**).

### Hospitalization due to COVID-19

mRNA-1273 was associated with a significantly lower risk of COVID-19–associated hospitalization versus BNT162b2 in the 9 studies included in the overall analysis (RR 0.83, 95% CI 0.76–0.90; *P*<0.0001; **Figure 2**). The RD (95% CI) of mRNA-1273 compared with BNT162b2 was estimated to be 60 fewer hospitalizations due to COVID-19 (from 140 fewer to 20 more). No evidence of heterogeneity was observed between studies (*I^2^*= 0%). The certainty of evidence for this outcome was type 3 (low) due to inclusion of nonrandomized studies and imprecision (**Table 2; Table S3**).

In 2 studies reporting hospitalization in patients with cancer (18; 45), mRNA-1273 was associated with a significantly reduced risk of hospitalization compared with BNT162b2 (RR 0.54, 95% CI 0.37–0.79; *P*=0.0013; RD −585, 95% CI −1,655 to 485). No association between mRNA vaccine type and COVID-19–associated hospitalization was found for the 3 studies each reporting hospitalization in the subgroups of patients with autoimmune diseases (RR 0.97, 95% CI 0.70–1.35; RD −103, 95% CI −1,661 to 1,456) (18; 71; 73) or solid organ transplant (RR 0.91, 95% CI 0.78–1.06; RD −147, 95% CI −816 to 522) (45; 46; 68). No evidence of heterogeneity was observed between any of the studies for the subgroup analysis. The certainty of evidence in all subgroups was graded as type 3 (low; **Table 3**).

### Death due to COVID-19

Of the 4 studies reporting COVID-19–associated mortality (46; 57; 66; 67), mRNA-1273 was associated with a significantly reduced risk of death compared with BNT162b2 (RR 0.62, 95% CI 0.43–0.89; *P*=0.011; **Figure 2**). mRNA-1273 was estimated to lead to 56 fewer deaths associated with COVID-19 (from 559 fewer to 446 more) compared with BNT162b2. No evidence of heterogeneity was observed between any of the studies (*I^2^*=0%). The certainty of evidence was rated as type 3 (low) due to inclusion of nonrandomized studies (**Table 2; Table S3**). Grading was reduced for imprecision and increased due to the strong association in RR.

COVID-19–associated death was assessed only in the subgroup of solid organ transplant recipients (46; 57; 67). In these 3 studies, mRNA-1273 was associated with a significantly reduced risk of death compared with BNT162b2 (RR 0.56, 95% CI 0.37–0.84; *P*=0.0049; RD −3,528, 95% CI −12,002 to 4,945). No evidence of heterogeneity was observed between any of the studies in this subgroup (*I^2^*=0%). The certainty of evidence was type 3 (low) due to inclusion of nonrandomized studies as well as imprecision and limited evidence (**Table 3**).

## Discussion

In this systematic review and pairwise meta-analysis of 22 studies, we found that mRNA-1273 was associated with a significantly lower risk of SARS-CoV-2 infection, hospitalization due to COVID-19, and COVID-19–associated mortality compared with BNT162b2 in adults with a broad spectrum of severe immunocompromising conditions. The certainty of this evidence was type 4 (very low) for the SARS-CoV-2 infection outcome and type 3 (low) for the COVID-19– associated hospitalization and death outcomes (**Table 4**). As all included studies were pairwise comparisons between mRNA-1273 and BNT162b2, the research question was not biased by differences in time period assessed, population, viral variants within each study. When outcomes were assessed in subgroups, mRNA-1273 was associated with significantly lower risk of SARS-CoV-2 infection and COVID-19–associated hospitalization versus BNT162b2 in patients with cancer. Compared with BNT162b2, mRNA-1273 was also associated with a significantly reduced risk of SARS-CoV-2 infection in patients with autoimmune diseases and COVID-19–associated death in solid organ transplant recipients.

IC individuals have a high burden of COVID-19 due to characteristics of their underlying disease or immunosuppressive treatments that impact their ability to mount productive immune responses as well as increased susceptibility to severe COVID-19 (30). Physicians may seek to optimize COVID-19 vaccine type, timing, and number of doses to improve outcomes in IC patients (32). RCTs are ranked highly in the hierarchy of evidence; however, studying comparative efficacy with adequate power would require enrolling a prohibitive number of IC patients. Therefore, the research question can only be assessed using large real-world databases where individual medical and pharmacy information is available.

Limitations of this systematic literature review were that non-English studies were excluded, and publication bias was not assessed in the meta-analysis. Inherent to the GRADE framework, evidence certainty is initially set to either high if the included studies are randomized studies or low if they are observational studies. As all but 1 of the 22 studies included in the pairwise meta-analysis were nonrandomized, the maximum certainty of evidence achievable in this meta-analysis was low despite the high number of observational studies and consistency of results. The pairwise meta-analysis was also limited by inconsistent outcome definitions across studies as well as differences in covariates between studies. For example, the vaccination scheme (2 vs 3 doses; booster) differed between studies, with a mix of primary series (100 mcg vs 30 mcg) and booster (50 mcg vs 30 mcg) pairwise comparisons included in the meta-analysis. Variants of concern changed over time, with risks of hospitalization and death (74) and vaccine effectiveness differing by variant (75). Vaccine effectiveness of 2-dose regimens could only be shown for the delta variant, whereas the omicron variant required a 3-dose schedule. Other sources of bias inherent to observational studies, such as prescribing differences by risk of severe COVID-19 and ability of patients to choose the mRNA vaccine type, could not be accounted for in this meta-analysis. In addition to differences in mRNA dosage between mRNA-1273 and BNT162b2, other differences such as the lipid nanoparticle delivery system and mRNA translation efficiency may also have impacted clinical effectiveness between vaccines.

Our meta-analysis of observational studies showed that mRNA-1273 (50 or 100 mcg/dose) was associated with a significantly reduced risk of SARS-CoV-2 infection, COVID-19-associated hospitalization, and death due to COVID-19 when compared with BNT162b2 (30 mcg/dose) in IC populations. Based on the findings, vaccinating IC individuals in the United States with mRNA-1273 instead of BNT162b2 would prevent an additional 60 and 56 hospitalizations and deaths per 100,000 individuals, respectively. Considering the limited availability of data from RCTs and to provide needed clinical decision-making guidance, our results showed that mRNA-1273 offers better clinical outcomes compared with BNT162b2 in vulnerable IC populations.

## Author Contributions

XW and KH designed and performed the systematic literature review and meta-analysis, and critically evaluated the manuscript. AS, LEP, AR, and AK designed and performed the systematic literature review and critically evaluated the manuscript. PS and SV collected data and critically evaluated the manuscript. MTB-J and NVdV conceptualized the article and provided oversight and critical evaluation of the manuscript. All authors contributed to the article and approved the submitted version.

## Data Availability

All data produced in the present work are contained in the manuscript.

## Acknowledgments

Writing assistance was provided by Erin McClure, PhD, an employee of ICON (Blue Bell, PA, USA) in accordance with Good Publication Practice (GPP3) guidelines, funded by Moderna, Inc., and under the direction of the authors.

## SUPPLEMENTAL MATERIALS

**Table S1.** Databases and Strategies Used for the Systematic Literature Review.

|  |  |  |
| --- | --- | --- |
| **Database** | WHO COVID-19 Global literature on coronavirus disease including MEDLINE (PubMed) |  |
| **URL** | https://search.bvsalud.org/global-literature-on-novel-coronavirus-2019-ncov/ |  |
| **No** | **Query scope** | **Query script** |
| 1 | All COVID-19 | Not applicable |
| 2 | Major focus on COVID-19 vaccine(s) or vaccination | mj: Covid-19 vacc* |
| 3 | Vaccine efficacy, effectiveness, related research types and terms | ("vaccine effectiveness" OR "vaccine efficacy" OR "vaccine research" OR "vaccine study" OR "vaccine trial" OR "vaccine control" ~2 OR "vaccine comparison"~2 OR "vaccine response" OR "IgM" OR "IgG" OR Ig* OR antibod* OR "anti-body" OR immunoglob* OR "immuno-globulin" OR immunogen* OR "immuno-genicity" OR sero* OR "immune response"~2) |
| 4 | At-risk medical conditions | ("risk condition"~2 OR "risk disorder"~2 OR "at-risk" ~2 OR "at risk" ~2 OR " high risk" OR "high-risk" OR "risk disease" ~2 OR "risk comorbidity" OR "risk disability" OR "long term condition"~2 OR "long-term condition"~2 OR underlying* OR pre-existing* OR preexisting* OR "medical condition" OR comorbid* OR immunocompr* OR immunodef* OR immunosupp* OR "immuno-compromised" OR "immuno-deficiency" OR "immuno-suppression" OR cancer* OR carcinoma* OR malignan* OR neoplasm* OR "solid organ" OR chemotherap* OR antineoplastic OR "anti-neoplastic" OR "cytotoxic therapy"~2 OR "anti-cancer" OR anticancer OR transplant* OR "stem cell" OR "SCT" OR "HSCT" OR "BMT" OR "chronic kidney" OR "CKD" OR "chronic liver" OR "CLD" OR cirrho* OR "chronic hepatitis" ~2 OR "HCC" OR haemochromatosis OR hemochromatosis OR "metabolic liver" OR "genetic liver" ~2 OR "chronic nephrotic"~2 OR "chronic |
|  |  | hepatic"~2 OR asthma* OR "bronchiectasis" OR "bronchopulmonary dysplasia" OR "chronic lung" OR "chronic pulmonary" OR "chronic obstructive" OR "COPD" OR emphysema OR "chronic bronchitis" OR "interstitial lung disease" OR "idiopathic pulmonary fibrosis" OR "pulmonary embolism" OR "pulmonary hypertension" OR "cystic fibrosis" OR diabet* OR "IDDM" OR "NIDDM" OR "overweight" OR obes* OR dementia OR Alzheimer* OR "degenerative brain"~2 OR "degenerative mental"~2 OR neurolog* OR neurodegen* OR schizo* OR "psychosis" OR "psychotic" OR "severe mood disorder" OR "clinical depression" OR "substance use" OR "substance abuse" OR "attention-deficit/hyperactivity disorder" OR "attention disorder"~2 OR "ADHD" OR "spinal cord injury" OR "cerebral palsy" OR "birth defect" ~2 OR "birth anomaly" OR "intellectual and developmental disability" OR "IDD" OR "learning disability" OR "Down syndrome" OR "inborn error" OR congenital OR inborn OR "inherited abnormality" OR "inherited disorder" OR "genetic abnormality"~2 OR "genetic disorder" ~2 OR heart OR cardiac OR coronary OR cardio* OR "CHD" OR "high blood pressure" OR "hypertension" OR myocard* OR cardiovasc* OR "HIV" OR "AIDS" OR "inherited red blood cell disorders" ~3 OR haemoglobinopath* OR hemoglobinopath* OR hemolytic OR haemolytic OR hematologic* OR haematologic* OR "stroke" OR "cerebrovascular" OR "CVD" OR "tuberculosis" OR "TB") |
| 5 | Without filtering for vaccine efficacy etc.<br>(#2 AND #4)<br>Major focus on COVID-19 | (mj: Covid-19 vacc*) AND ("risk condition"~2 OR "risk disorder"~2 OR "at-risk" ~2 OR "at risk" ~2 OR " high risk" OR "high-risk" OR "risk disease" ~2 OR "risk comorbidity" OR "risk disability" OR "long term condition"~2 OR "long-term condition"~2 OR underlying* OR pre-existing* OR preexisting* OR "medical |
|  | vaccine(s) or vaccination<br>AND<br>At-risk medical conditions | condition" OR comorbid* OR immunocompr* OR immunodef* OR immunosupp*<br>OR "immuno-compromised" OR "immuno-deficiency" OR "immuno-suppression"<br>OR cancer* OR carcinoma* OR malignan* OR neoplasm* OR "solid organ" OR<br>chemotherap* OR antineoplastic OR "anti-neoplastic" OR "cytotoxic therapy"~2<br>OR "anti-cancer" OR anticancer OR transplant* OR "stem cell" OR "SCT" OR<br>"HSCT" OR "BMT" OR "chronic kidney" OR "CKD" OR "chronic liver" OR "CLD"<br>OR cirrho* OR "chronic hepatitis" ~2 OR "HCC" OR haemochromatosis OR<br>hemochromatosis OR "metabolic liver" OR "genetic liver" ~2 OR "chronic<br>nephrotic"~2 OR "chronic hepatic"~2 OR asthma* OR "bronchiectasis" OR<br>"bronchopulmonary dysplasia" OR "chronic lung" OR "chronic pulmonary" OR<br>"chronic obstructive" OR "COPD" OR emphysema OR "chronic bronchitis" OR<br>"interstitial lung disease" OR "idiopathic pulmonary fibrosis" OR "pulmonary<br>embolism" OR "pulmonary hypertension" OR "cystic fibrosis" OR diabet* OR<br>"IDDM" OR "NIDDM" OR "overweight" OR obes* OR dementia OR Alzheimer*<br>OR "degenerative brain"~2 OR "degenerative mental"~2 OR neurolog* OR<br>neurodegen* OR schizo* OR "psychosis" OR "psychotic" OR "severe mood<br>disorder" OR "clinical depression" OR "substance use" OR "substance abuse"<br>OR "attention-deficit/hyperactivity disorder" OR "attention disorder"~2 OR<br>"ADHD" OR "spinal cord injury" OR "cerebral palsy" OR "birth defect" ~2 OR<br>"birth anomaly" OR "intellectual and developmental disability" OR "IDD" OR<br>"learning disability" OR "Down syndrome" OR "inborn error" OR congenital OR<br>inborn OR "inherited abnormality" OR "inherited disorder" OR "genetic<br>abnormality"~2 OR "genetic disorder" ~2 OR heart OR cardiac OR coronary OR<br>cardio* OR "CHD" OR "high blood pressure" OR "hypertension" OR myocard* |
|  |  | OR cardiovasc* OR "HIV" OR "AIDS" OR "inherited red blood cell disorders" ~3<br>OR haemoglobinopath* OR hemoglobinopath* OR hemolytic OR haemolytic OR<br>hematologic* OR haematologic* OR "stroke" OR "cerebrovascular" OR "CVD" OR<br>"tuberculosis" OR "TB") |
| 6 | **Full query**<br>**(#2 AND #3 AND #4)**<br>**Major focus on COVID-19**<br>**vaccine(s) or vaccination AND**<br>**Vaccine efficacy, effectiveness,**<br>**related research types and**<br>**terms AND**<br>**At-risk medical conditions** | (mj: Covid-19 vacc*) AND ("vaccine effectiveness" OR "vaccine efficacy" OR<br>"vaccine research" OR "vaccine study" OR "vaccine trial" OR "vaccine control" ~2<br>OR "vaccine comparison"~2 OR "vaccine response" OR "IgM" OR "IgG" OR Ig*<br>OR antibod* OR "anti-body" OR immunoglob* OR "immuno-globulin" OR<br>immunogen* OR "immuno-genicity" OR sero* OR "immune response"~2) AND<br>("risk condition"~2 OR "risk disorder"~2 OR "at-risk" ~2 OR "at risk" ~2 OR " high<br>risk" OR "high-risk" OR "risk disease" ~2 OR "risk comorbidity" OR "risk disability"<br>OR "long term condition"~2 OR "long-term condition"~2 OR underlying* OR pre-<br>existing* OR preexisting* OR "medical condition" OR comorbid* OR<br>immunocompr* OR immunodef* OR immunosupp* OR "immuno-compromised"<br>OR "immuno-deficiency" OR "immuno-suppression" OR cancer* OR carcinoma*<br>OR malignan* OR neoplasm* OR "solid organ" OR chemotherap* OR<br>antineoplastic OR "anti-neoplastic" OR "cytotoxic therapy"~2 OR "anti-cancer"<br>OR anticancer OR transplant* OR "stem cell" OR "SCT" OR "HSCT" OR "BMT"<br>OR "chronic kidney" OR "CKD" OR "chronic liver" OR "CLD" OR cirrho* OR<br>"chronic hepatitis" ~2 OR "HCC" OR haemochromatosis OR hemochromatosis<br>OR "metabolic liver" OR "genetic liver" ~2 OR "chronic nephrotic"~2 OR "chronic<br>hepatic"~2 OR asthma* OR "bronchiectasis" OR "bronchopulmonary dysplasia"<br>OR "chronic lung" OR "chronic pulmonary" OR "chronic obstructive" OR "COPD"<br>OR emphysema OR "chronic bronchitis" OR "interstitial lung disease" OR |
|  |  | "idiopathic pulmonary fibrosis" OR "pulmonary embolism" OR "pulmonary hypertension" OR "cystic fibrosis" OR diabet* OR "IDDM" OR "NIDDM" OR "overweight" OR obes* OR dementia OR Alzheimer* OR "degenerative brain"~2 OR "degenerative mental"~2 OR neurolog* OR neurodegen* OR schizo* OR "psychosis" OR "psychotic" OR "severe mood disorder" OR "clinical depression" OR "substance use" OR "substance abuse" OR "attention-deficit/hyperactivity disorder" OR "attention disorder"~2 OR "ADHD" OR "spinal cord injury" OR "cerebral palsy" OR "birth defect" ~2 OR "birth anomaly" OR "intellectual and developmental disability" OR "IDD" OR "learning disability" OR "Down syndrome" OR "inborn error" OR congenital OR inborn OR "inherited abnormality" OR "inherited disorder" OR "genetic abnormality"~2 OR "genetic disorder" ~2 OR heart OR cardiac OR coronary OR cardio* OR "CHD" OR "high blood pressure" OR "hypertension" OR myocard* OR cardiovasc* OR "HIV" OR "AIDS" OR "inherited red blood cell disorders" ~3 OR haemoglobinopath* OR hemoglobinopath* OR hemolytic OR haemolytic OR hematologic* OR haematologic* OR "stroke" OR "cerebrovascular" OR "CVD" OR "tuberculosis" OR "TB") |
| 7 | **Full query**<br>**(#2 AND #3 AND #4)**<br>**Major focus on COVID-19 vaccine(s) or vaccination AND Vaccine efficacy, effectiveness, related research types and terms AND** | (mj: covid-19 vacc*) AND ("vaccine effectiveness" OR "vaccine efficacy" OR "vaccine research" OR "vaccine study" OR "vaccine trial" OR "vaccine control" ~2 OR "vaccine comparison"~2 OR "vaccine response" OR "IgM" OR "IgG" OR ig* OR antibod* OR "anti-body" OR immunoglob* OR "immuno-globulin" OR immunogen* OR "immuno-genicity" OR sero* OR "immune response"~2) AND ("risk condition"~2 OR "risk disorder"~2 OR "at-risk" ~2 OR "at risk" ~2 OR " high risk" OR "high-risk" OR "risk disease" ~2 OR "risk comorbidity" OR "risk disability" |
|  | **At-risk medical conditions**<br>**English publications** | OR "long term condition"~2 OR "long-term condition"~2 OR underlying* OR pre-existing* OR preexisting* OR "medical condition" OR comorbid* OR immunocompr* OR immunodef* OR immunosupp* OR "immuno-compromised" OR "immuno-deficiency" OR "immuno-suppression" OR cancer* OR carcinoma* OR malignan* OR neoplasm* OR "solid organ" OR chemotherap* OR antineoplastic OR "anti-neoplastic" OR "cytotoxic therapy"~2 OR "anti-cancer" OR anticancer OR transplant* OR "stem cell" OR "SCT" OR "HSCT" OR "BMT" OR "chronic kidney" OR "CKD" OR "chronic liver" OR "CLD" OR cirrho* OR "chronic hepatitis" ~2 OR "HCC" OR haemochromatosis OR hemochromatosis OR "metabolic liver" OR "genetic liver" ~2 OR "chronic nephrotic"~2 OR "chronic hepatic"~2 OR asthma* OR "bronchiectasis" OR "bronchopulmonary dysplasia" OR "chronic lung" OR "chronic pulmonary" OR "chronic obstructive" OR "COPD" OR emphysema OR "chronic bronchitis" OR "interstitial lung disease" OR "idiopathic pulmonary fibrosis" OR "pulmonary embolism" OR "pulmonary hypertension" OR "cystic fibrosis" OR diabet* OR "IDDM" OR "NIDDM" OR "overweight" OR obes* OR dementia OR alzheimer* OR "degenerative brain"~2 OR "degenerative mental"~2 OR neurolog* OR neurodegen* OR schizo* OR "psychosis" OR "psychotic" OR "severe mood disorder" OR "clinical depression" OR "substance use" OR "substance abuse" OR "attention-deficit/hyperactivity disorder" OR "attention disorder"~2 OR "ADHD" OR "spinal cord injury" OR "cerebral palsy" OR "birth defect" ~2 OR "birth anomaly" OR "intellectual and developmental disability" OR "IDD" OR "learning disability" OR "Down syndrome" OR "inborn error" OR congenital OR inborn OR "inherited abnormality" OR "inherited disorder" OR "genetic abnormality"~2 OR "genetic disorder" ~2 OR |
|  |  | heart OR cardiac OR coronary OR cardio* OR "CHD" OR "high blood pressure" OR "hypertension" OR myocard* OR cardiovasc* OR "HIV" OR "AIDS" OR "inherited red blood cell disorders" ~3 OR haemoglobinopath* OR hemoglobinopath* OR hemolytic OR haemolytic OR hematologic* OR haematologic* OR "stroke" OR "cerebrovascular" OR "CVD" OR "tuberculosis" OR "TB") AND la:("en") |
| **Database** | WHO COVID-19 Global literature on coronavirus disease excluding MEDLINE (Pubmed); including other reference sources in the WHO COVID-19 database (ICTRP; EMBASE; EuropePMC; PREPRINT-MEDRXIV; Web of Science; ProQuest Central; Academic Search Complete; Scopus; COVIDWHO)– full query #3 |  |
| **URL** | https://search.bvsalud.org/global-literature-on-novel-coronavirus-2019-ncov/ |  |
| **No** | **Query scope** | **Query script** |
| 1 | All COVID-19 | Not applicable |
| 2 | WHO accessed databases other than MEDLINE (PubMed) | db:("ICTRP" OR "EMBASE" OR "EuropePMC" OR "PREPRINT-MEDRXIV" OR "Web of Science" OR "ProQuest Central" OR "Academic Search Complete" OR "Scopus" OR "COVIDWHO") |
| 3 | COVID-19 vaccine(s) or vaccination AND vaccine efficacy, effectiveness, related research types and terms AND At-risk medical conditions AND WHO accessed databases other than MEDLINE (PubMed) | ("covid19 vaccine" ~2 OR "covid-19 vaccine"~2 OR "covid-19 vaccines"~2 OR "covid19 vaccines" ~2 OR "covid-19 vaccination"~2 OR "Covid19 vaccination"~2) AND ("vaccine effectiveness" OR "vaccine efficacy" OR "vaccine research" OR "vaccine study" OR "vaccine trial" OR "vaccine control" ~2 OR "vaccine comparison"~2 OR "vaccine response" OR "IgM" OR "IgG" OR Ig* OR antibod* OR "anti-body" OR immunoglob* OR "immuno-globulin" OR immunogen* OR "immuno-genicity" OR sero* OR "immune response"~2) AND ("risk condition"~2 OR "risk disorder"~2 OR "risk disease" ~2 OR "risk comorbidity" OR "risk disability" or "long term condition"~2 OR "long-term condition"~2 OR underlying* |
|  |  | OR pre-existing* OR preexisting* OR "medical condition" OR comorbid* OR immunocompr* OR immunodef* or immunosupp* OR "immuno-compromised" OR "immuno-deficiency" OR "immuno-suppression" or cancer* OR carcinoma* OR malignan* OR neoplasm* OR "solid organ" OR chemotherap* OR antineoplastic OR "anti-neoplastic" OR "cytotoxic therapy"~2 OR "anti-cancer" OR anticancer or transplant* OR "stem cell" OR "SCT" OR "HSCT" OR "BMT" OR "chronic kidney" OR "CKD" OR "chronic liver" OR "CLD" OR cirrho* OR "chronic hepatitis" ~2 OR "HCC" OR haemochromatosis OR hemochromatosis or "metabolic liver" OR "genetic liver" ~2 OR "chronic nephrotic"~2 OR "chronic hepatic"~2 OR asthma* OR "bronchiectasis" OR "bronchopulmonary dysplasia" OR "chronic lung" OR "chronic pulmonary" or "chronic obstructive" OR "COPD" OR emphysema OR "chronic bronchitis" OR "interstitial lung disease" OR "idiopathic pulmonary fibrosis" OR "pulmonary embolism" OR "pulmonary hypertension" OR "cystic fibrosis" OR diabet* OR "IDDM" OR "NIDDM" OR "overweight" OR obes* OR dementia OR alzheimer* OR "degenerative mental"~2 OR neurolog* OR neurodegen* OR schizo* or "psychosis" OR "psychotic" OR "severe mood disorder" OR "clinical depression" OR "substance use" OR "substance abuse" OR "attention-deficit/hyperactivity disorder" OR "attention disorder"~2 or "ADHD" OR "spinal cord injury" OR "cerebral palsy" OR "birth defect" ~2 OR "birth anomaly" OR "intellectual and developmental disability" OR "IDD" OR "learning disability" OR "Down syndrome" or "inborn error" OR congenital OR inborn OR "inherited abnormality" OR "inherited disorder" OR "genetic abnormality"~2 OR "genetic disorder" ~2 OR heart OR cardiac OR coronary OR cardio* OR "CHD" or "high blood pressure" OR "hypertension" OR myocard* OR cardiovasc* OR "HIV" |
|  |  | OR "AIDS" OR "inherited red blood cell disorders" ~3 OR haemoglobinopath* OR<br>hemoglobinopath* OR hemolytic OR haemolytic OR hematologic* OR<br>haematologic* OR "stroke" OR "cerebrovascular" OR "CVD" OR "tuberculosis"<br>OR "TB") AND db:("ICTRP" OR "EMBASE" OR "EuropePMC" OR "PREPRINT-<br>MEDRXIV" OR "Web of Science" OR "ProQuest Central" OR "Academic Search<br>Complete" OR "Scopus" OR "COVIDWHO") |

**Table S2.** Research Question and PICOS.

|  |  |  |
| --- | --- | --- |
| **Research question** | Is the mRNA-1273 COVID-19 vaccine (50 or 100 mcg/dose) more clinically effective in IC populations compared with the BNT162b2 COVID-19 vaccine (30 mcg/dose)? |  |
|  | **Include** | **Exclude** |
| **Population** | IC individuals $\geq 18$ years of age defined as people with CEV groups 1 and 2 medical conditions (51) | • Pregnant women, current/former smokers, physically inactive<br>• Studies on only healthy individuals or individuals not categorized as CEV |
| **Intervention** | mRNA-1273 | Studies with heterologous vaccination schedule (i.e., data on mixed mRNA-1273, BNT162b2, or other vaccines) |
| **Comparison** | BNT162b2 |  |
| **Outcomes** | • Vaccine efficacy/effectiveness against COVID-19 infection<br>• Vaccine efficacy/effectiveness against symptomatic COVID-19<br>• Vaccine efficacy/effectiveness against severe COVID-19<br>• Vaccine efficacy/effectiveness against hospitalization<br>• Vaccine efficacy/effectiveness against death<br>• SARS-CoV2 positivity (symptomatic or asymptomatic)<br>• Symptomatic laboratory-confirmed COVID-19 infection<br>• Severe COVID-19 infection (hospitalization or death)<br>• Breakthrough infection<br>• Hospitalization due to COVID-19 (ICU, ER, or ventilation etc.) | Studies only with safety results |
|  | • Death due to COVID-19 |  |
| **Study design** | • Clinical trials<br>• Observational studies<br>• Any kind of real-world evidence | • Study protocol (no results)<br>• Economic models |
| **Other limits** | • Any publication type (including letters, commentary, abstract, full text, poster)<br>• Publication in English |  |

CEV, clinically extremely vulnerable; COVID-19, coronavirus disease 2019; ER, emergency room; IC, immunocompromised; ICU, intensive care unit; PICOS, population, intervention, comparison, outcomes, and study design; SARS-CoV-2, severe acute respiratory syndrome coronavirus 2; SLR, systematic literature review.

**Table S3.**
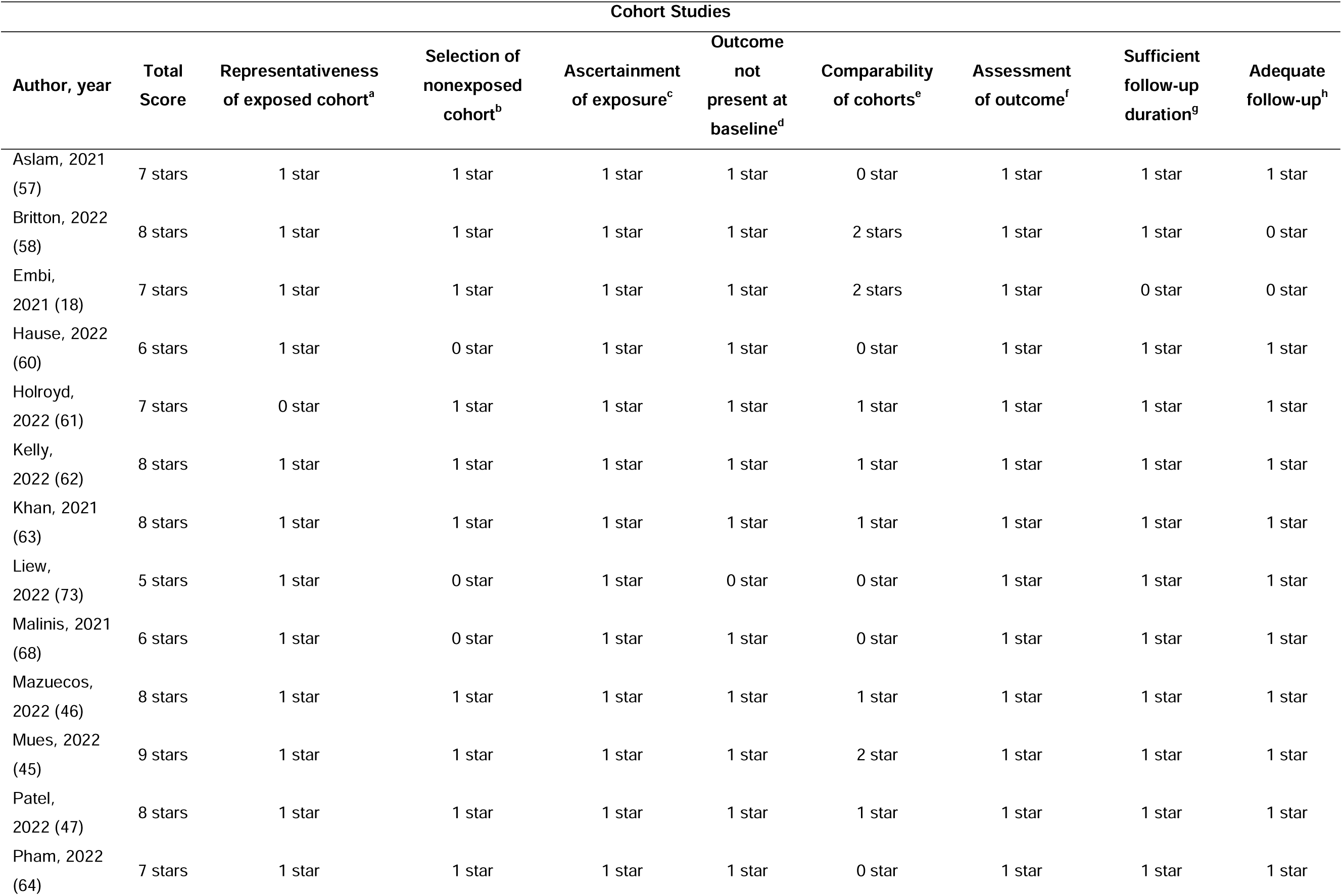
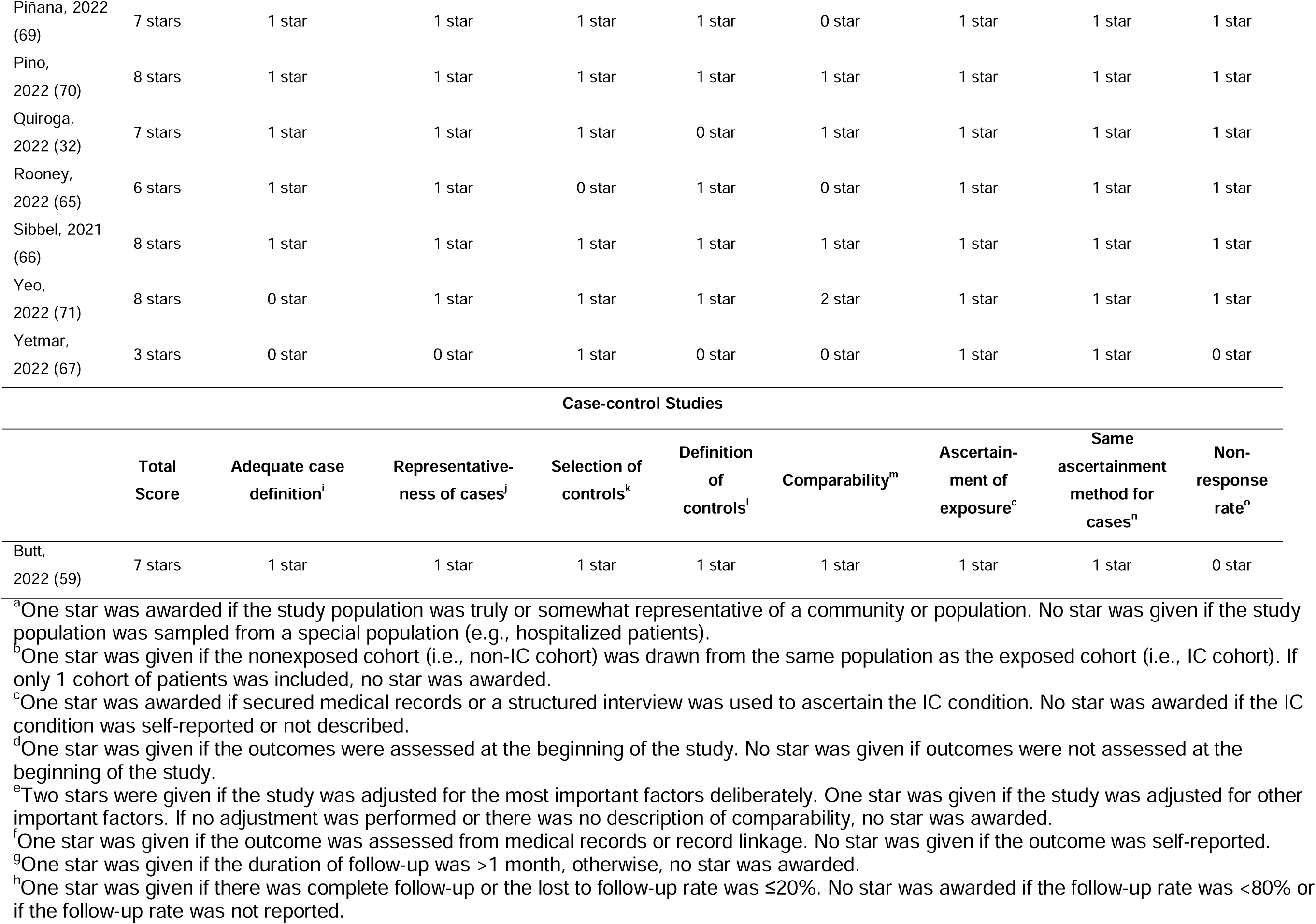
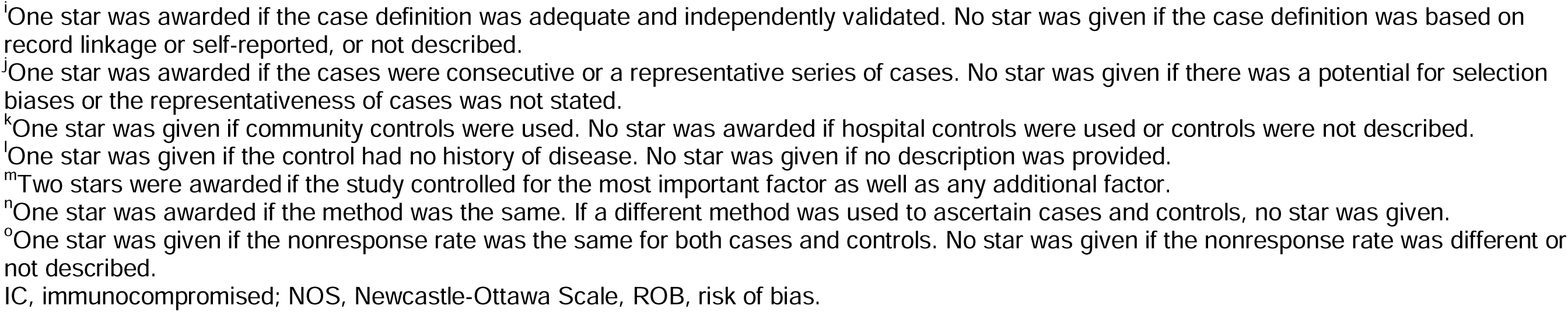
ROB Assessment per the NOS Scale for Cohort and Case-Control Studies.

**Table S4.**
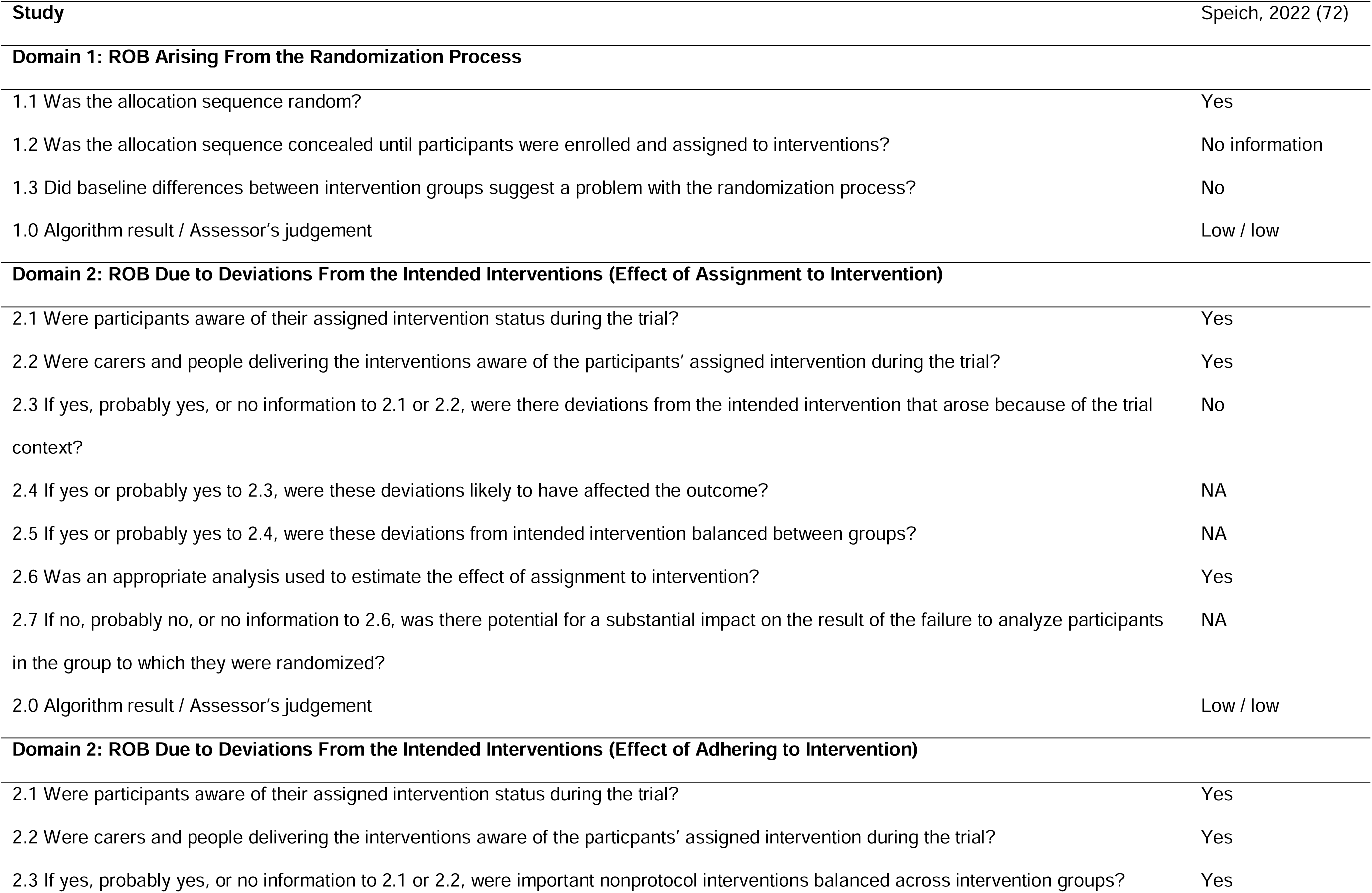
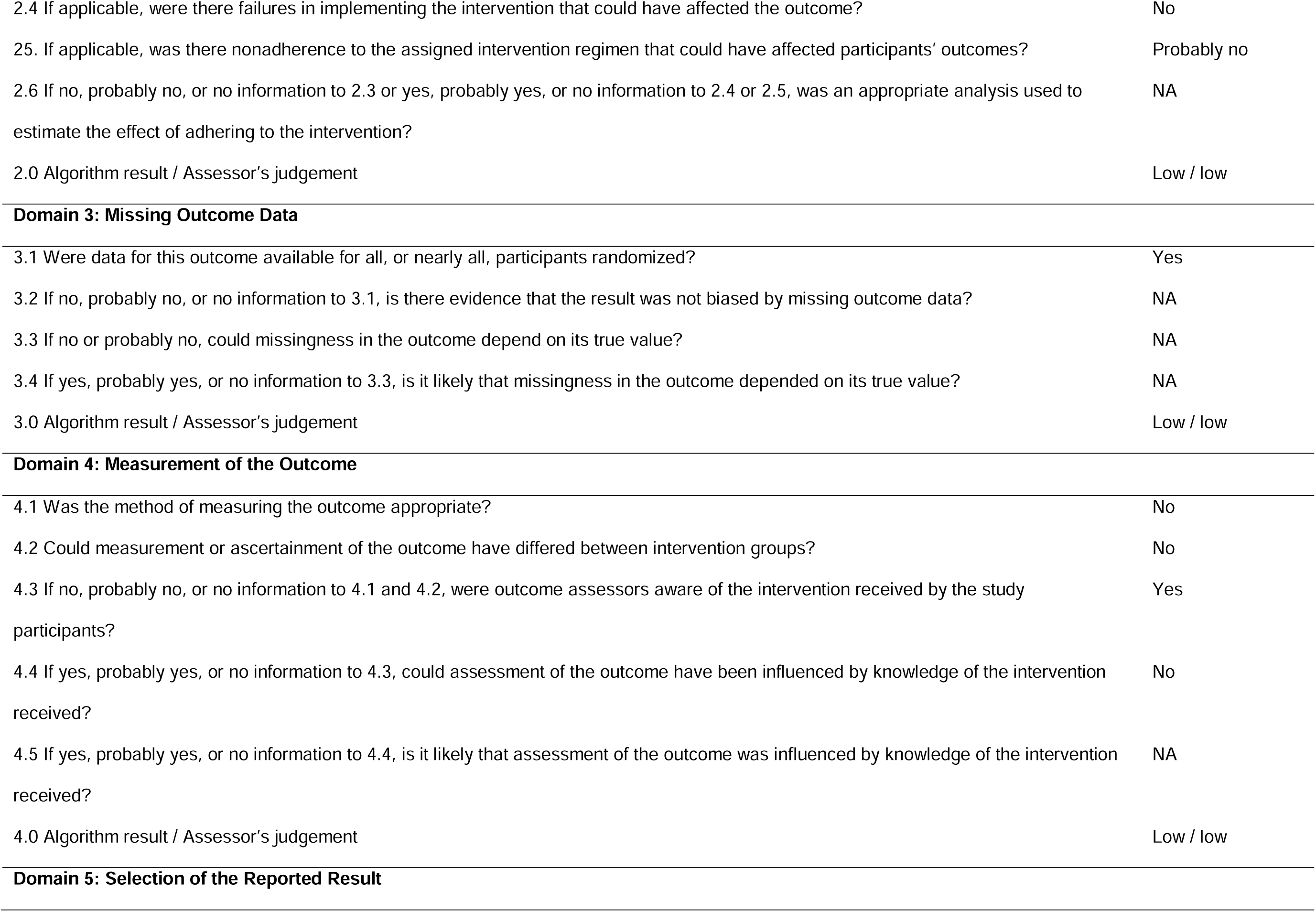
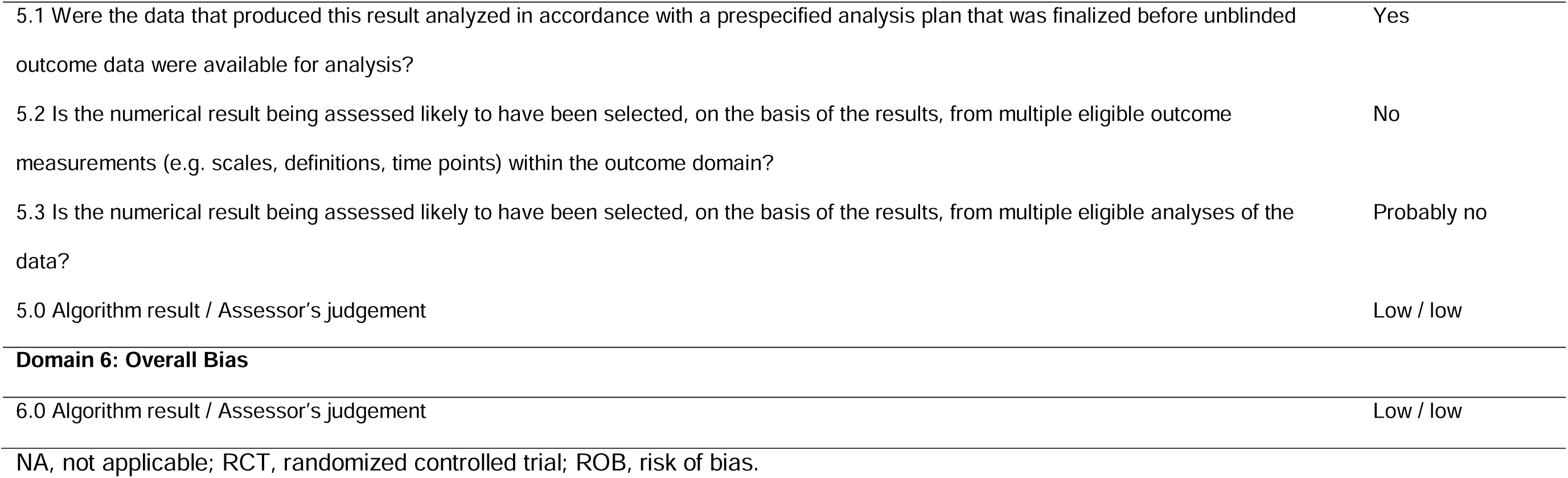
ROB Assessment for RCTs.

|  |  |
| --- | --- |
| **Study** | Speich, 2022 (72) |
| **Domain 1: ROB Arising From the Randomization Process** |  |
| 1.1 Was the allocation sequence random? | Yes |
| 1.2 Was the allocation sequence concealed until participants were enrolled and assigned to interventions? | No information |
| 1.3 Did baseline differences between intervention groups suggest a problem with the randomization process? | No |
| 1.0 Algorithm result / Assessor's judgement | Low / low |
| **Domain 2: ROB Due to Deviations From the Intended Interventions (Effect of Assignment to Intervention)** |  |
| 2.1 Were participants aware of their assigned intervention status during the trial? | Yes |
| 2.2 Were carers and people delivering the interventions aware of the participants' assigned intervention during the trial? | Yes |
| 2.3 If yes, probably yes, or no information to 2.1 or 2.2, were there deviations from the intended intervention that arose because of the trial context? | No |
| 2.4 If yes or probably yes to 2.3, were these deviations likely to have affected the outcome? | NA |
| 2.5 If yes or probably yes to 2.4, were these deviations from intended intervention balanced between groups? | NA |
| 2.6 Was an appropriate analysis used to estimate the effect of assignment to intervention? | Yes |
| 2.7 If no, probably no, or no information to 2.6, was there potential for a substantial impact on the result of the failure to analyze participants in the group to which they were randomized? | NA |
| 2.0 Algorithm result / Assessor's judgement | Low / low |
| **Domain 2: ROB Due to Deviations From the Intended Interventions (Effect of Adhering to Intervention)** |  |
| 2.1 Were participants aware of their assigned intervention status during the trial? | Yes |
| 2.2 Were carers and people delivering the interventions aware of the participants' assigned intervention during the trial? | Yes |
| 2.3 If yes, probably yes, or no information to 2.1 or 2.2, were important nonprotocol interventions balanced across intervention groups? | Yes |
| 2.4 If applicable, were there failures in implementing the intervention that could have affected the outcome? | No |
| 25. If applicable, was there nonadherence to the assigned intervention regimen that could have affected participants' outcomes? | Probably no |
| 2.6 If no, probably no, or no information to 2.3 or yes, probably yes, or no information to 2.4 or 2.5, was an appropriate analysis used to estimate the effect of adhering to the intervention? | NA |
| 2.0 Algorithm result / Assessor's judgement | Low / low |
| Domain 3: Missing Outcome Data |  |
| 3.1 Were data for this outcome available for all, or nearly all, participants randomized? | Yes |
| 3.2 If no, probably no, or no information to 3.1, is there evidence that the result was not biased by missing outcome data? | NA |
| 3.3 If no or probably no, could missingness in the outcome depend on its true value? | NA |
| 3.4 If yes, probably yes, or no information to 3.3, is it likely that missingness in the outcome depended on its true value? | NA |
| 3.0 Algorithm result / Assessor's judgement | Low / low |
| Domain 4: Measurement of the Outcome |  |
| 4.1 Was the method of measuring the outcome appropriate? | No |
| 4.2 Could measurement or ascertainment of the outcome have differed between intervention groups? | No |
| 4.3 If no, probably no, or no information to 4.1 and 4.2, were outcome assessors aware of the intervention received by the study participants? | Yes |
| 4.4 If yes, probably yes, or no information to 4.3, could assessment of the outcome have been influenced by knowledge of the intervention received? | No |
| 4.5 If yes, probably yes, or no information to 4.4, is it likely that assessment of the outcome was influenced by knowledge of the intervention received? | NA |
| 4.0 Algorithm result / Assessor's judgement | Low / low |
| Domain 5: Selection of the Reported Result |  |
| 5.1 Were the data that produced this result analyzed in accordance with a prespecified analysis plan that was finalized before unblinded outcome data were available for analysis? | Yes |
| 5.2 Is the numerical result being assessed likely to have been selected, on the basis of the results, from multiple eligible outcome measurements (e.g. scales, definitions, time points) within the outcome domain? | No |
| 5.3 Is the numerical result being assessed likely to have been selected, on the basis of the results, from multiple eligible analyses of the data? | Probably no |
| 5.0 Algorithm result / Assessor's judgement | Low / low |
| **Domain 6: Overall Bias** |  |
| 6.0 Algorithm result / Assessor's judgement | Low / low |
| NA, not applicable; RCT, randomized controlled trial; ROB, risk of bias. |  |

